# The association of child marriage with morbidities and mortality among children under 5 years in Afghanistan: Findings from a national survey

**DOI:** 10.1101/2022.06.09.22276186

**Authors:** Omid Dadras, Mohammadsediq Hazratzai, Fateme Dadras

**Author notes:** **Corresponding author:** Omid Dadras, Department of Global Public Health and Primary Care, University of Bergen, Norway.

## Abstract

**Background:** There is still a gap in knowledge of the impact that child marriage could have on the mortality and morbidity of children in Afghanistan. This study used the data from the latest Afghanistan demographic health survey conducted in 2015 (ADHS) to address this gap and advance the current knowledge.

**Methods:** A secondary analysis of the 2015 ADHS, including the births in the past 5 years to ever-married women aged 15-24 years old, was carried out. Logistic regression analyses were employed to examine the association of child marriage (<18y) with morbidities (diarrhea, acute respiratory infection, and fever in the last 2 weeks), mortality (neonatal, infant, child), and size at birth among the children under 5 born to women aged 15-24 years, before and after adjusting for the effect of sociodemographic and structural inequalities.

**Results:** Approximately two-thirds of births in the past 5 years belong to 15-24 years old mothers who married at ages <18. The majority of them were born to mothers residing in rural areas (75.67%) with no education (51.68%) from poor households (39.39%). As compared to the births to women married at ages ≥18, there was a significantly higher likelihood of neonatal mortality among births to women married at ages <18 (crude OR = 2.30, 95% CI: 1.52-3.49 & adjusted OR = 1.94, 95% CI: 1.25-3.01) and higher infant mortality among the births to the women married at ages ≤14y (crude OR = 1.94, 95% CI: 1.06-3.53). However, it disappeared for neonatal mortality after adjustment for adequacy of antenatal care (ANC) and infant mortality after adjustment for sociodemographic inequalities.

**Conclusion:** Although the births to women married as a child (<18) were more likely to die at an early age, this association disappeared after adjustment for the adequacy of ANC. Given the unavoidable practice of child marriage in Afghanistan, this finding emphasizes the importance of providing adequate ANC for young brides to prevent child mortality. In addition, strong global advocacy is required to empower and support young Afghan women in negotiating their reproductive and maternity rights with their partners by reducing social and gender-based inequalities.

## Introduction

Child marriage is described as marriage before the age of 18 years. It is a major social problem, mainly in low- and middle-income countries (LMICs), associated with several adverse health outcomes in young women (1). Moreover, it is also a violation of human rights and a serious public health issue (2). Although there have been enormous efforts to reduce child marriage worldwide during the last decades, this harmful practice remained prevalent in some regions such as Sub-Saharan Africa (38%), South Asia (30%), and Latin America (25%) (3). Additionally; in some Asian Muslim countries such as Afghanistan and Pakistan, the influence of strict Islamic culture caused such practices to be more common; for example in 2018, more than a third of women were married before age of 18 years in Pakistan (4) and Afghanistan (5).

The Afghan Civil Law allows marriage at ages ≥16 years for girls and ≥18 years for boys; however, in Afghan culture, younger marriage even at very early ages is encouraged especially for girls. Moreover, the widespread poverty in Afghanistan encourages poor families to force their young girls into marriage at very early ages in hope of a better life and receiving large doweries from future husbands (6). Child marriage is also a common practice for settling debts and making peace with rivals in Afghanistan (7). That being said, child marriage has been a barrier to gender equality efforts, and the achievement of sustainable development goals targeted specifically goals 1 to 5: The eradication of poverty, the achievement of universal education, women empowerment, reducing child mortality, and the improvement of maternal health (8).

Marriage at early ages (<18 years) has been linked to several adverse maternal and child outcomes. It has been shown that child brides are at a higher risk for unintended and high-risk pregnancy, sexually transmitted diseases, and obstetric fistula (9-11). Moreover, there are more likely to suffer from intimate partner violence and subsequent depression and this could further harm maternal and child health (6). A variety of sociocultural factors such as lack of education, limited access to media, insufficient health literacy, and man-dominant norms have been described as potential drivers for such trends (12-14). In addition, a lack of power in decision-making and negotiating reproductive rights could lead to the inability to actively engage in health promotion programs and adequately receive the necessary health care during pregnancy which could lead to adverse outcomes (15-17). Nevertheless, it has been shown that maternity and reproductive health programs lack the necessities to address these obstacles and are; moreover, hard to reach these vulnerable populations in LMICs (2, 18). This could not only endanger the mother’s health but also the child’s health.

Early marriage has also been linked to low birth weight and higher child and infant morbidity and mortality in some LIMCs (19-21). However, the results are inconsistent across countries and more epidemiological data is required to formulate well-tailored interventions targeting the child brides to ensure the health of both child mothers and offspring. In addition, there is still a gap in knowledge of the true associations of age at marriage with mortality and morbidity among children in some of the most afflicted countries such as Afghanistan. Thus, this study aimed to explore the impact that child marriage could have on the morbidity and mortality of children, taking into account the effect of social and structural inequalities, among a nationally representative sample of children under 5 years in Afghanistan. The results could inform and guide future policies and interventions to enhance the health of Afghan child brides and their offspring.

## Methods

### Study design

This cross-sectional study analyzed the data for births in the past 5 years in the latest Afghanistan Demographic and Health Survey (ADHS) carried out in 2015. ADHS is a nationally representative survey implemented by the Central Statistics Organization (CSO) in collaboration with the Afghanistan Ministry of Public Health (MoPH) and funded by the United States Agency for International Development (USAID).

### Study population and sampling

ADHS 2015 collected data for women aged 15-49 years and their children aged less than 5 years old. A stratified two-stage cluster sampling was employed to estimate the key indicators at the national level, in urban and rural areas, and for each of the 34 provinces of Afghanistan. In the first stage, 950 clusters□enumeration areas from the previous national census□including 260 urban and 690 rural areas were selected. In the second stage, through an equal probability systematic selection process, 25,650 households were selected. Sampling weights were calculated and applied to obtain a representative estimate at the national level. All women aged 15-49 years (n=29641) who were either permanent residents of the selected households or visitors who stayed in the households the night before the survey were recruited after informed consent (22). The women were interviewed about all births in the past 5 years (n = 31063). Our analyses were restricted to the births of young women aged 15-24 to ensure the inclusion of all the births in the last 5 years to the mothers that married at early ages (<18). Including the births to this maternal age group not only enhances the comparability of the results across similar studies but also reduces the chance of recall bias which may occur as the women move to the older age groups (23). Moreover, we could obtain estimates that reflect relatively recent marriages rather than marriages that occurred decades ago.

### Measurement and scales

A standard DHS questionnaire collected the data related to the demographic and health of all eligible women and their children under 5 including women’s demographic characteristics, household information, family planning, fertility preferences, child health, sexually transmitted diseases, marriage, sexual behavior, and domestic violence (24). However, for the current study, our primary interest was questions related to child marriage and morbidity and mortality indicators of children under 5 years.

### Independent variables

The main explanatory variable was maternal age at first marriage and child marriage was defined as marriage at ages less than 18 years for the first time. The age at marriage was further categorized into three groups: ≥18 years (marriage as an adult), 15-17 years (marriage in middle adolescence), and ≤14 years of age (marriage in early adolescence or childhood) to capture the potential vulnerability that may exist for the birth to the mothers married in very early ages.

### Outcomes of interest

In total, eight indicators concerning the morbidity and mortality of children under 5 years were estimated and their relationship with maternal age at marriage was examined. These included:

#### Morbidity indicators

- ***Diarrhea in the past 2 weeks***: This was assessed by a question if the child had diarrhea within the last two weeks and coded as “1=yes” and “0=no”.
- ***Acute respiratory infection (ARI) in the past 2 weeks***: This was assessed by two questions (ie, whether the child had suffered from a cough and rapid breathing in the last two weeks) and coded as “1=yes” and “0=no”.
- ***Fever in the past two weeks***: This was assessed by questions about whether the child had fever in the last two weeks and coded as “1=yes” and “0=no”.
- ***Very small or small size at birth***: This was reported by the mother as the size of the child at birth, was dichotomized into “very small or smaller than average” and “average or larger”.

#### Mortality indicators

- ***Child mortality:*** This was defined as the death of the child before the age of 5 years (0-59 months).
- ***Infant mortality***: This was defined as death within the first year of life.
- ***Neonatal mortality***: This was defined as death within the first month of life.

### Covariates

Covariates included the sociodemographic characteristics of births to young women aged 15-24 years consisting of maternal and child factors. Maternal demographic included age, education, place of residence, wealth index, ethnicity, maternal age at birth, and the adequacy of antenatal care (ANC) visits (< 4 times, ≥4 based on the WHO recommendation for optimal ANC visits). Child-specific demographics included age, sex, and being born to a multiparous birth.

### Statistical Analysis

All the analyses were restricted to the births of ever-married women aged 15-24 years in the last 5 years. The descriptive statistics described the maternal and child demographics and compared them across the categories of maternal age at marriage (<18 years vs ≥18 years) using the Chi-square test (categorical variables) and t-student test (continuous variables) (Table 1). Logistic regression analysis, applying the sampling design to account for the clustering of multiple births within one mother at the household level, was employed to estimate the odds of the child morbidity and mortality indices across the categories of maternal age at marriage (<18 years vs ≥18 years); before and after adjustment for maternal and child’s sociodemographic factors (Table 2). Adjusted models for diarrhea, ARI, and fever outcomes included only the living children under 5-year and accounted for the age of child, sex of child, multiparous births, maternal age, maternal education, place of residence, and wealth index. Adjusted models for child mortality indices, birth size, and low birth weight outcomes included all the births in the past 5 years and accounted for sex of child, multiparous births, maternal age, maternal education, place of residence, and wealth index. The birth order and father’s education were dropped in the final multivariate models due to multicollinearity issues that existed between these variables with the mother’s age and education. In addition to the mentioned covariates, a separate model was built to account for the impact of antenatal visits to examine the effect of age at marriage on child morbidity and mortality indices beyond the attributable influence of sociodemographic inequalities, including the inadequate number of antenatal visits during pregnancy. Given the current evidence indicating lower age at marriage could increase the risk of maternal and child morbidity and mortality (25, 26), additional models were constructed to compare the child morbidity and mortality indices across different categories of maternal age at marriage (≤14 years, 15-17, ≥18). Results were presented as odds ratio (OR) and adjusted odds ratio (AOR) with 95% confidence intervals (95% CI). All the analyses were performed using STATA version 14. The sampling weight and design were counted for using the STATA command “svy” in all analyses. A 2-tailed P value <.05 was considered to be statistically significant.

**Table 1.**
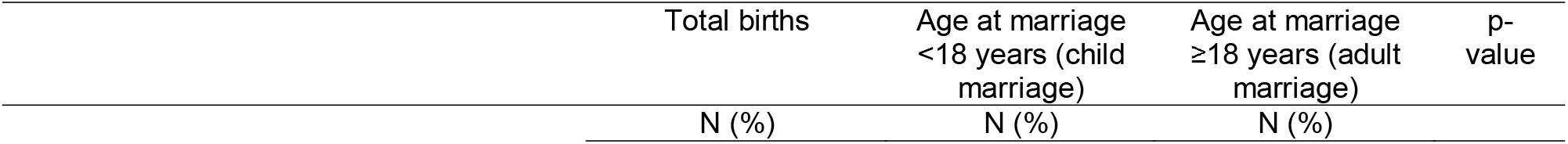
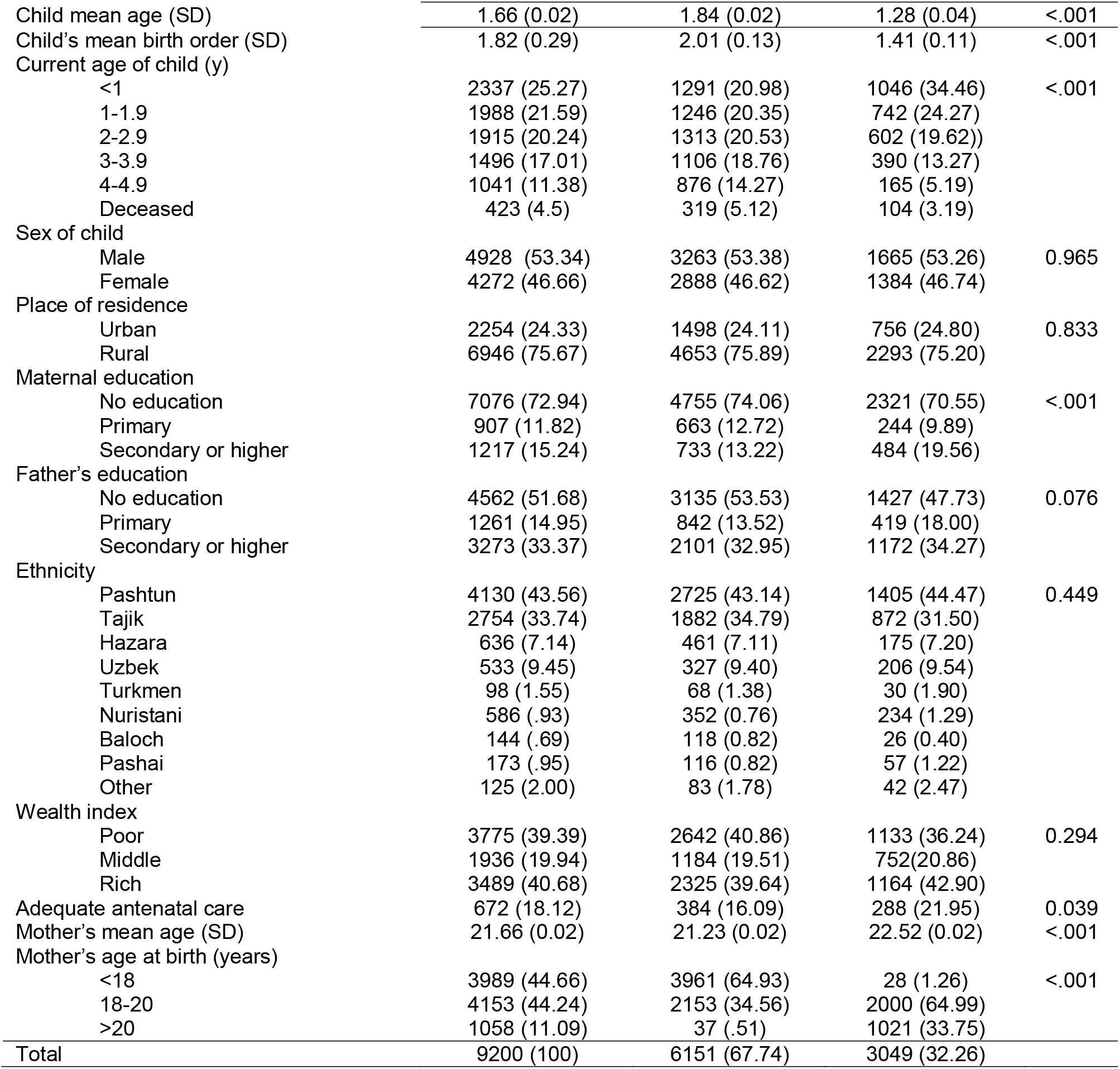
Demographics of births in the past 5 years to ever-married women aged 15-24 years in Afghanistan, by maternal age at marriage

| Total births | Age at marriage<br><18 years (child<br>marriage) | Age at marriage<br>$\geq 18$ years (adult<br>marriage) | p-value |
| --- | --- | --- | --- |
| N (%) | N (%) | N (%) |  |

|  |  |  |  |  |
| --- | --- | --- | --- | --- |
| Child mean age (SD) | 1.66 (0.02) | 1.84 (0.02) | 1.28 (0.04) | <.001 |
| Child's mean birth order (SD) | 1.82 (0.29) | 2.01 (0.13) | 1.41 (0.11) | <.001 |
| Current age of child (y) |  |  |  |  |
| <1 | 2337 (25.27) | 1291 (20.98) | 1046 (34.46) | <.001 |
| 1-1.9 | 1988 (21.59) | 1246 (20.35) | 742 (24.27) |  |
| 2-2.9 | 1915 (20.24) | 1313 (20.53) | 602 (19.62) |  |
| 3-3.9 | 1496 (17.01) | 1106 (18.76) | 390 (13.27) |  |
| 4-4.9 | 1041 (11.38) | 876 (14.27) | 165 (5.19) |  |
| Deceased | 423 (4.5) | 319 (5.12) | 104 (3.19) |  |
| Sex of child |  |  |  |  |
| Male | 4928 (53.34) | 3263 (53.38) | 1665 (53.26) | 0.965 |
| Female | 4272 (46.66) | 2888 (46.62) | 1384 (46.74) |  |
| Place of residence |  |  |  |  |
| Urban | 2254 (24.33) | 1498 (24.11) | 756 (24.80) | 0.833 |
| Rural | 6946 (75.67) | 4653 (75.89) | 2293 (75.20) |  |
| Maternal education |  |  |  |  |
| No education | 7076 (72.94) | 4755 (74.06) | 2321 (70.55) | <.001 |
| Primary | 907 (11.82) | 663 (12.72) | 244 (9.89) |  |
| Secondary or higher | 1217 (15.24) | 733 (13.22) | 484 (19.56) |  |
| Father's education |  |  |  |  |
| No education | 4562 (51.68) | 3135 (53.53) | 1427 (47.73) | 0.076 |
| Primary | 1261 (14.95) | 842 (13.52) | 419 (18.00) |  |
| Secondary or higher | 3273 (33.37) | 2101 (32.95) | 1172 (34.27) |  |
| Ethnicity |  |  |  |  |
| Pashtun | 4130 (43.56) | 2725 (43.14) | 1405 (44.47) | 0.449 |
| Tajik | 2754 (33.74) | 1882 (34.79) | 872 (31.50) |  |
| Hazara | 636 (7.14) | 461 (7.11) | 175 (7.20) |  |
| Uzbek | 533 (9.45) | 327 (9.40) | 206 (9.54) |  |
| Turkmen | 98 (1.55) | 68 (1.38) | 30 (1.90) |  |
| Nuristani | 586 (.93) | 352 (0.76) | 234 (1.29) |  |
| Baloch | 144 (.69) | 118 (0.82) | 26 (0.40) |  |
| Pashai | 173 (.95) | 116 (0.82) | 57 (1.22) |  |
| Other | 125 (2.00) | 83 (1.78) | 42 (2.47) |  |
| Wealth index |  |  |  |  |
| Poor | 3775 (39.39) | 2642 (40.86) | 1133 (36.24) | 0.294 |
| Middle | 1936 (19.94) | 1184 (19.51) | 752 (20.86) |  |
| Rich | 3489 (40.68) | 2325 (39.64) | 1164 (42.90) |  |
| Adequate antenatal care | 672 (18.12) | 384 (16.09) | 288 (21.95) | 0.039 |
| Mother's mean age (SD) | 21.66 (0.02) | 21.23 (0.02) | 22.52 (0.02) | <.001 |
| Mother's age at birth (years) |  |  |  |  |
| <18 | 3989 (44.66) | 3961 (64.93) | 28 (1.26) | <.001 |
| 18-20 | 4153 (44.24) | 2153 (34.56) | 2000 (64.99) |  |
| >20 | 1058 (11.09) | 37 (.51) | 1021 (33.75) |  |
| Total | 9200 (100) | 6151 (67.74) | 3049 (32.26) |  |

**Table 2.**
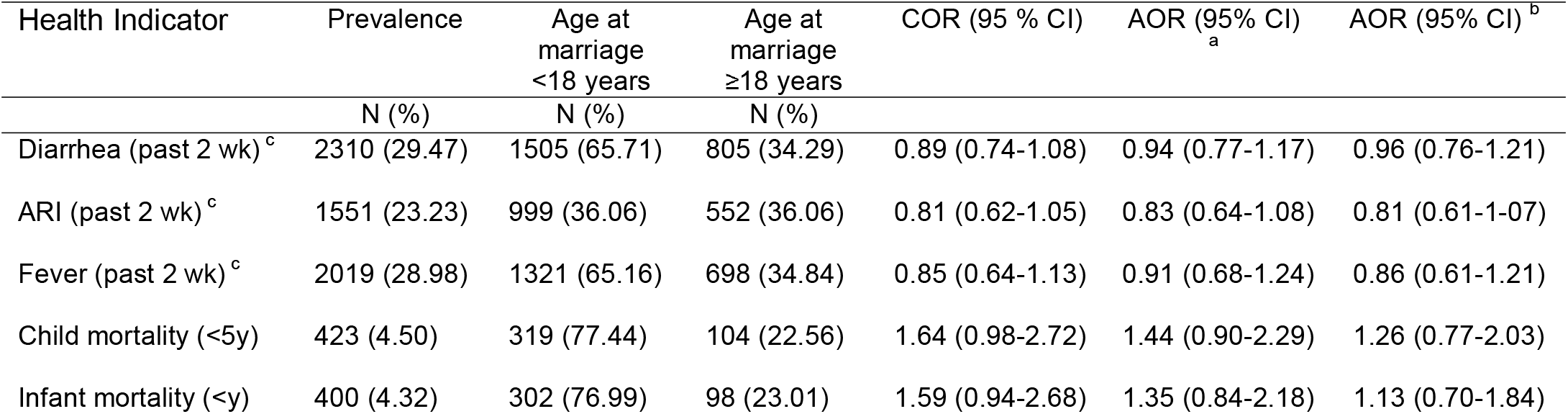
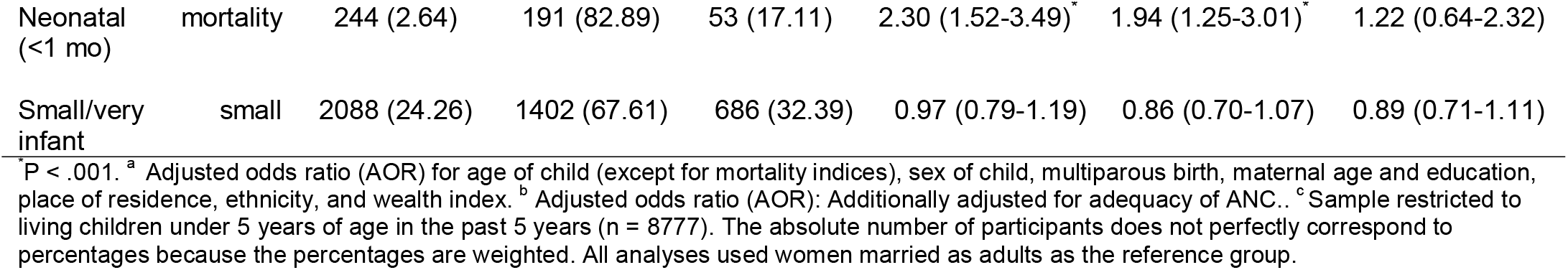
The association of maternal child marriage with morbidity, mortality, and low birth weight among births in the past 5 years to ever-married women aged 15-24 years in Afghanistan

## Results

### 1. Sociodemographic of the children under 5 years old, ADHS 2015

A total of 9200 births to ever-married women aged 15-24 years old were included in the final analysis; of whom, 6151 (67.74%) were born to women married as a child (<18 years) and 3049 (32.26%) were the births to women married as an adult (≥18 years). The mean age of the children was significantly higher among the births to women married as a child (1.84 yrs ± 0.02 in child marriage vs 1.28 yrs ± 0.04 in the adult marriage group; p<0.001). Likewise, as is shown in Table 1, the proportions of children in older age groups were higher among the births to women married as a child as compared to those born to women married as an adult. More than half the births were male in both child and adult marriage groups. The place of residence was not significantly different between the births of women married as a child compared to counterparts and in both groups only a quarter of children were residing in urban areas (Table 1). Approximately two-thirds of women had no education; however, the rate of no education was **Table 1**. Demographics of births in the past 5 years to ever-married women aged 15-24 years in Afghanistan, by maternal age at marriage significantly higher among the women married as a child compared to those married as an adult (74.06% vs 70.55%, p<0.001). Similarly, the rate of no education was higher among the fathers in the child marriage group in comparison with those in the adult marriage group (53.33% vs 47.73%); however, not significant (p=0.076). The majority of the children under 5 were from Pashtun and Tajik ethnic groups (43.56% and 33.74%, respectively). In terms of the wealth index, there were no significant differences between births to women of either group (child vs adult marriage) and almost equal proportions of poor and rich in both groups. Only 18.2% of women aged 15-24 years had adequate ANC in their recent pregnancy and the rate of adequate ANC was significantly higher among the adult marriage group as compared to the child marriage group (21.95% versus 16.09%, p-value=0.039). The mean age of the mother was 21.23 yrs ± 0.02 in the child marriage group; which was significantly lower than the adult marriage group (22.52 yrs ± 0.02, p<0.001). As expected, the majority of the births to women married at ages <18 occurred at younger ages (64.93% of mothers were <18 yrs old or younger at the time of birth) as compared to the births to women married at ages ≥18 (64.99% mothers were 18-20 yrs old at the time of birth).

### 2. The prevalence of morbidities and mortality indices among children under 5 and their association with maternal age at marriage

The prevalence of diarrhea, ARI, and fever in the last 2 weeks was respectively 29.47%, 23.23%, and 28.98% among children under 5 years. There were no significant differences between the rate of diarrhea, ARI, and fever in the last 2 weeks between the children born to women married at ages <18 and those born to women married at ages ≥18 (Table 2).

The child, infant, and neonatal mortality rates were respectively 4.5% (423), 4.32% (400), and 2.34% (244). Except for the neonatal mortality rate (COR = 2.30, 95% CI: 1.52-3.49 and AOR = 1.94, 95% CI: 1.25-3.01), the child and infant mortality rates were not statistically different between the babies born to the women married at ages <18 and those born to women married at ages ≥18. However, the difference in neonatal mortality rate became insignificant after adjustment for the adequacy of ANC. Furthermore; in the extended age-at-marriage analysis (≤14, 15-17, ≥18), the difference in infant mortality was significant among births to women married before age 14 as compared to births to women married at older ages (COR = 1.94, 95% CI: 1.06-3.53) in unadjusted model and turned to be insignificant in the adjuted model (Tables 2 and 3).

**Table 3.** The association of maternal age at marriage with morbidity, mortality, and low birth weight among births in the past 5 years to ever-married women aged 15-24 years in Afghanistan, by marital age group

| Health Indicator | COR (95 % CI) | AOR (95%CI)<sup>a</sup> | AOR (95%CI)<sup>b</sup> |
| --- | --- | --- | --- |
| Diarrhea (past 2 wk)<sup>c</sup> |  |  |  |
| ≤14 | 0.91 (0.74-1.11) | 0.95 (0.73-1.25) | 0.96 (0.70-1.31) |
| 15-17 | 0.85 (0.67-1.09) | 0.94 (0.76-1.17) | 0.95 (0.74-1.21) |
| 18≥ | Reference | Reference | Reference |
| ARI (past 2 wk) <sup>c</sup> |  |  |  |
| ≤14 | 0.74 (0.53-1.04) | 0.77 (0.55-1.08) | 0.70 (0.49-1.02) |
| 15-17 | 0.83 (0.63-1.09) | 0.84 (0.64-1.11) | 0.84 (0.63-1.11) |
| 18≥ | Reference | Reference | Reference |
| Fever (past 2 wk) <sup>c</sup> |  |  |  |
| ≤14 | 0.76 (0.56-1.03) | 0.84 (0.62-1.15) | 0.71 (0.50-1.03) |
| 15-17 | 0.88 (0.64-1.21) | 0.94 (0.68-1.31) | 0.91 (0.63-1.30) |
| 18≥ | Reference | Reference | Reference |
| Child mortality (<5y) |  |  |  |
| ≤14 | 1.98 (1.10-3.56) | 1.34 (0.83-2.19) | 1.73 (0.91-3.29) |
| 15-17 | 1.52 (0.90-2.57) | 1.75 (0.99-3.10) | 1.11 (0.66-1.88) |
| 18≥ | Reference | Reference | Reference |
| Infant mortality (<y) |  |  |  |
| ≤14 | 1.94 (1.06-3.53) <sup>a</sup> | 1.67 (0.94-2.96) | 1.58 (0.82-3.06) |
| 15-17 | 1.48 (0.87-2.53) | 1.27 (0.78-2.06) | 0.99 (0.59-1.68) |
| 18≥ | Reference | Reference | Reference |
| Neonatal mortality (<1 mo) |  |  |  |
| ≤14 | 2.74 (1.57-4.80) <sup>a</sup> | 2.33 (1.27-4.27) <sup>a</sup> | 1.69 (0.73-3.92) |
| 15-17 | 2.16 (1.40-3.34) <sup>a</sup> | 1.82 (1.16-2.86) <sup>a</sup> | 1.07 (0.54-2.12) |
| 18≥ | Reference | Reference | Reference |
| Small/very small infant |  |  |  |
| ≤14 | 1.12 (0.87-1.45) | 0.96 (0.74-1.23) | 0.87 (0.68-1.11) |
| 15-17 | 0.92 (0.75-1.14) | 0.83 (0.66-1.05) | 0.96 (0.73-1.28) |
| 18≥ | Reference | Reference | Reference |

<sup>a</sup> P < .001.
<sup>a</sup> Adjusted odds ratio (AOR) for age of child (except for mortality indices), sex of child, multiparous birth, maternal age and education, place of residence, ethnicity, and wealth index.
<sup>b</sup> Adjusted odds ratio (AOR): Additionally adjusted for adequacy of ANC.
<sup>c</sup> Sample restricted to living children under 5 years of age in the past 5 years (n = 8777).

Approximately a quarter of births were described as very small or small in size at the time of birth by their mother. It appeared that there is no difference in the rate of very small or small size at birth with maternal age at marriage even after adjustment for sociodemographic factors and the adequacy of ANC (Tables 2 and 3).

## Discussion

To the best of our knowledge, this was the first study that assess the prevalence and association of child marriage with morbidities, mortality, and birth size among children under 5 born to ever-married women aged 15-24 years old in Afghanistan using the data from lastest ADHS conducted in 2015. The results showed that almost two-thirds of births to women aged 15-24 were born to mothers married as a child (<18 years). The majority of these births were born to mothers from socially disadvantaged backgrounds (i.e lower education, living in rural areas, poor families) from Pashtun and Tajik ethnicities. Higher poverty and illiteracy documented in these ethnic groups could be moderators for marriage at very early ages (27). Similar findings were documented in other Asian countries with high rates of child marriages such as Pakistan (28), Bangladesh (29), and India (9). The births to women married as a child (<18) were more likely to die at an early age; however, this association disappeared after adjustment for the adequacy of ANC; indicative of the important role of adequate ANC in preventing premature death of newborns.

Child marriage is common practice in Afghan society and has been a rampant exercise for decades to strengthen the ties between rival families and tribes and settle the conflict and debts among them. Girls in poor families are often forced to marry a man, usually much older than them, in hope of a better life and in exchange for large dowries (7, 27). Additionally, the recent conflict and transfer of power in Afghanistan have pushed more Afghan families to the brink of poverty and exposed more families and children to hunger (6). The strict fanatic civil laws that have been recently imposed by the Taliban are an obvious violation of women’s rights and facilitate forced child marriage in Afghanistan which could lead to irreparable health outcomes for both young Afghan women and their offspring. In the current situation, strong global advocacy and initiatives are necessary to mitigate the burden of early and forced marriage for Afghan girls and the threat of immature death and morbidities among newborns.

The threat that child marriage could impose on neonatal and child health has been documented in several countries; however, the results are inconsistent; for instance, a study among the births in the past 5 years to ever-married Pakistani women aged 15-24 years observed higher child (<5 yrs) and infant (<1yr) mortality, as well as a higher prevalence of the diarrheal diseases in unadjusted models among the births to the women, married at ages <18 years (28). On the contrary; in our study, except for neonatal and infant mortality, other morbidities and mortality indices including child mortality, recent diarrhea, ARI, and fever were not significantly different between the children born to women married as a child (<18) as compared to those born to women married as adults (≥18yrs). This could be due to contextual and demographic diversity as well as the methodological differences between these two studies. The distinctive feature of our study was accounting for the clustering effect that exists among children of a single mother that could produce more accurate and reliable standard errors. Another study among children under 5 who were born to women aged 15-24 years in India (9) also found contradictory results, reporting a higher rate of diarrheal diseases among the children born to women married as a child compared to those born to women married as an adult (9, 28).

The higher mortality and morbidity among the births of child mothers have been attributed to the lower volume of breast milk that a physically immature mother could produce and possibly a lower concentration of colostrum which contains the necessary antibodies against infective microorganisms in early life (30, 31). It is also could be due to maternal nutrient depletion as a result of repeated pregnancies, particularly among poor and less educated young mothers (28). However, this association often disappeared after adjustment for social inequalities and the number of ANC visits in these studies. Likewise, the association of neonatal and infant mortality with child marriage observed in our study disappeared after adjustment for the adequacy of ANC, this may implicate the lower number of ANC as the potential culprit for higher morbidity and mortality among the births to child mothers. It is well known that inadequate ANC could lead to a higher incidence of adverse reproductive and maternal outcomes (32, 33); and it is more likely for Afghan women married at ages <18 years, particularly those living in rural and poor urban areas, to have inadequate maternity care partially due to the lack of power in decision-making when it comes to negotiating their maternal and reproductive needs with their husbands (27). That being said, we argue that it is not only the physical immaturity of the young brides but also their inadequate access to maternity care that could cause higher morbidity and mortality among their children. However, more in-depth longitudinal data are required to ascertain such an association. Nonetheless, scaling maternity care services through leveraging the existent capacity at both individual and community levels is strongly recommended in order to enhance the access of the child mothers to such services and improve the health outcomes of both young mothers and their offspring.

## Limitations

Although this study offers valuable insights into child marriage prevalence and its association with morbidity and mortality among children under 5 years old in Afghanistan; The results should be interpreted in light of some limitations. First, the majority of outcomes were self-reported; thus, there is a risk of socially desirable response and recall bias. However, we restricted our analysis to the births of women aged 15-24 years to reduce the rate of recall bias by including the recent marriage and births rather than those of decades ago. Therefore, the findings are not representative of all the births in the past five years but merely of those born to the mother aged 15-24 years old. In addition, the results are specific to the young women in Afghanistan and are not generalizable to other settings and contexts. Another limitation was the cross-sectional nature of the data that hinder the assumption of causality; however, as all marriage precedes birth in the primary analysis, the ordering of this exposure relative to child health outcomes could stand correct. However, longitudinal data are necessary to precisely ascertain the effect that maternal age at marriage could have on neonatal and child health outcomes. Lastly, the lack of anthropometrical data such as weight and height in ADHS 2015 did not allow us to estimate the indices related to growth retardation such as stunting and wasting that have been linked to child marriage in previous studies (9).

## Conclusion

In summary, the results indicated higher neonatal mortality among the births to women married at ages <18 years. Additionally, those born to women married at ages ≤14 years were more likely to die before their first birthday (< 1 year). However, this association disappeared after adjustment for the adequacy of ANC; indicative of the important role of adequate ANC in preventing child mortality. The inadequate ANC is linked to several adverse pregnancy outcomes, particularly among child brides in Afghanistan, and it is mainly attributed to the lack of power in decision-making and negotiating their reproductive rights with partners (6). This also emphasizes the importance of women empowerment in receiving adequate maternity care and reducing child morbidity and mortality regardless of the mother’s age. Additionally, with the recent tragic turn of events in Afghanistan and the transfer of power to the Taliban, the obvious violation of young women’s rights is nothing to be easily ignored. Hence, strong global advocacy is needed to rectify the violation of young Afghan women’s rights and empower this vulnerable population; dictating appropriate interventions and policies to reduce the gender-based inequalities and consequent adverse health outcomes in both child mothers and offspring. Some solutions could be increasing the age of marriage by maintaining adolescent girls at school, delaying childbirth by appropriate reproductive health education, and reducing socioeconomic and cultural vulnerabilities that may decrease cause-specific child morbidity and mortality in young adolescent mothers in Afghanistan. Nonetheless, further longitudinal research is necessary to ascertain the exact factors that influence child mortality and morbidity among child mothers.

## Data Availability

The DHS questionnaire that collected the data in Afghanistan's demographic and health survey in 2015 could be downloaded from DHS official website (https://www.dhsprogram.com). The dataset (ADHS 2015) that was used in this study could be available upon a reasonable request and with permission from either the Walailak University ethical board or the DHS website.

## Abbreviations

DHS: Demographic and Health Survey
EA: Enumeration area
ANC: Antenatal care
LMICs: low- and middle-income countries
COR: Crude odds ratio
AOR: Adjusted odds ratio

## Declarations

### Ethics approval and consent to participate

The study was performed in accordance with relevant guidelines and regulations (U.S. Department of Health and Human Services regulations for the protection of human subjects). In addition, this survey was approved by the Institutional Review Board (IRB) of the Afghanistan Ministry of Health (MoH). An informed verbal and written consent was obtained from all the participants/guardians/parents before the interview. We also sought permission from the DHS website and filled out a request to access and download the data. Therefore, further ethical approval to use the data is not necessary.

### Consent for publication

Not applicable.

## Availability of data and material

The DHS questionnaire that collected the data in Afghanistan’s demographic and health survey in 2015 could be downloaded from DHS’s official website (https://www.dhsprogram.com). The main dataset (ADHS 2015) that was used in this study could be available upon request by the registered user from the DHS website or by contacting the corresponding author of this manuscript.

## Authors’ contribution

OD, FD conceptualized the study and wrote the study protocol. OD and MH performed the data analysis and organized the tables and figures. OD wrote the first draft. OD, MH, and FD revised the first draft, provided critical comments, and revised the final draft. All authors read the final draft and agreed upon it before submission.

## Acknowledgment

We would like to express our utmost gratitude to the librarians at the library of the University of Bergen for providing technical support to prepare and submit this manuscript.

